# Comparative profiling of epigenetic modifications among individuals living in different high and low air pollution zones: A pilot study from India

**DOI:** 10.1101/2020.09.15.20194928

**Authors:** Pradyumna Kumar Mishra, Neha Bunkar, Radha Dutt Singh, Rajat Kumar, Pushpendra Kumar Gupta, Rajnarayan Tiwari, Lalit Lodhi, Arpit Bhargava, Koel Chaudhury

**Author notes:** **Address for Correspondence:** Prof. (Dr.) Pradyumna Kumar Mishra, MS, PhD, Fulbright-Nehru Fellow (USA), Dy. Director & Head, Department of Molecular Biology, ICMR-National Institute for Research in Environmental Health, Kamla Nehru Hospital Building (Gandhi Medical College Campus), Bhopal (MP)-462001, India; https://orcid.org/0000-0002-Q795-2819.

## Abstract

Epigenetic modifications act as an important bridge to regulate the complex network of gene-environment interaction. As these mechanisms determines the gene-expression patterns via regulating the transcriptomic machinery, environmental stress induced epigenetic modifications may interrupt distinct cellular functions resulting into generation of diseased phenotypes. In the present study, we used a multi-city approach to compare the epigenomic signatures of individuals living in two tiers of Indian cities categorized as low-risk and high-risk air pollution zones. The high-risk group reported marked changes in the expression levels of epigenetic modifiers (DNMT1, DNMT3a, EZH2, EHMT2 and HAT), that maintains the levels of specific epigenetic marks essential for appropriate gene functioning. These results also coincided with the observed alterations in the levels of DNA methylation (LINE-1 and % 5mC), and histone modifications (H3 and H4), among the high-risk group. In addition, higher degree of changes reported in the expression profile of a selected miRNA panel in the high-risk group indicated the probability of deregulated transcriptional machinery. This was further confirmed by the analysis of a target gene panel involved in various signalling pathways, which revealed differential expression of the gene transcripts regulating cell cycle, inflammation, cell survival, apoptosis and cell adhesion. Together, our results provide first insights of epigenetic modifications among individuals living in different high and low levels of air pollution zones of India. However, further steps to develop a point-of-care epigenomic assay for human bio-monitoring may be immensely beneficial to reduce the health burden of air pollution especially in lower-middle-income countries.

## 1. Introduction

Air pollution arising from a composite mixture of pollutants (derived from both natural and anthropogenic sources) is now considered as a serious health concern (Landrigan et al., 2018). The global burden of this exposure can be comprehended by the fact that in 2017, it was the 5^th^ major risk factor for mortality and was related to almost 4.9 million deaths and 147 million years of healthy life lost (SOGA, 2019). The particulate matter (PM) emitted from combustion sources or formed through atmospheric chemical transformation is one of the major contributors of air pollution. Although, World Health Organization (WHO) has set up interim targets (IT) for regulation of the daily and annual mean concentrations of PM (classified as class I carcinogen), more than 91% of the world’s population still lives in the regions exceeding WHO’s limits of healthy air (SOGA, 2019). Unfortunately, the seriousness of PM exposure in India is alarming, as 1 out of 8 deaths in the country in 2017 were attributed to the air pollution (Venkataraman et al., 2018; India State-Level Disease Burden Initiative Air Pollution Collaborators, 2019). The country ranked 5^th^ among most polluted countries in 2019 with an annual mean levels of PM_2.5_ reaching to 58.1 μg/m^3^. Among the list of world’s 50 most polluted cities in 2019, 26 cities (≥50%) were from India (World Air Quality Report, 2019). To tackle this, several programmes including National Clean Air Programme (NCAP) have been initiated, which aims to reduce the PM_2.5_ and PM_10_ concentration in 102 cities by 20-30% by 2024. In a country where no region falls below IT-3 levels (≤10 μg/m^3^), this national initiative may be highly beneficial as approximately 80% of the Indian population is exposed to PM levels much beyond WHO IT-1 norms (≤35 μg/m^3^). Almost 673,000 deaths and 21.3 million disability-adjusted life years were attributed to the long term ambient PM_2.5_ exposure (SOGA, 2019).

Of late, several interesting studies have shown that exposure to PM induces significant molecular repercussions in vital organ functions, affecting heart, lungs, and central nervous system (Yang et al., 2018; Shen et al., 2018; Chen et al., 2020). Majority of these effects were attributed to presence of different organic and metallic compounds that remain closely associated with PM and their capability to induce oxidative damage (Duan et al., 2016). Few important studies have delineated the plausible mechanistic insights of the involvement of various biological pathways (Lyu et al., 2018; Chen et al., 2018; Chu et al., 2019). Earlier we have shown that *in vitro* PM exposure to human lymphocytes not only disturbs the mitochondrial machinery and triggers PI-3-kinase mediated DNA damage response; but this impairment further activates NF-κβ associated inflammatory loop that has an important role in the regulation of downstream epigenetic mechanisms (Bhargava et al., 2018; Bhargava et al., 2019). As epigenetic mechanisms are dynamic in nature and can be profoundly influenced by the surrounding environmental factors, these mechanisms act as an essential interface between the environmental signals and regulation of genomic responses (Bunkar et al., 2016; Feinberg, 2018). The major interacting systems (DNA methylation, histone modifications and miRNA expressions) coordinate to ensure the proper regulation of biological processes and the somatically heritable states of gene expression. However, any disturbance in this tightly regulated machinery is likely to transform the gene functionality thereby resulting in different disease outcomes which could also be transferred vertically (Feinberg, 2018; Shukla et al., 2019). As the association DNA methylation (Li et al., 2019a; Eze et al., 2020); histone modifications (Zheng et al., 2017; Li et al., 2018); and miRNA expressions (Mancini et al., 2020; Cheng et al., 2020) with PM exposure is evident, we aimed to perform a comparative profiling of epigenetic modifications among individuals living in different high and low levels of air pollution zones from India.

## 2. Material and methods

### 2.1. Sample collection & processing

The study was conducted under the IMPRINT India Initiative programme funded by Ministry of Health and Family Welfare (MoHFW) and Ministry of Human Resource Development (MHRD), Government of India. Based on the air quality index (AQI; μg/m^3^), the cities were categorized as low-risk (Good; ≤50), and high-risk (Unhealthy for sensitive groups; ≥100) zones. The high-risk cities selected for the study were Delhi, Raipur and Gwalior, while Sagar, Nayagarh and Mandla were included as low-risk cities. Five mL of blood samples (via venipuncture method) from healthy and non-smoker individuals from each city (including 120 samples, 60 in each group) were collected during August 2017 to August 2018. Written consent with the details of age, marital status, year of residence, socio-economic status, education, occupation, family income, type of exposure such as outdoor and indoor exposure, and fuel type used for the cooking were recorded appropriately. Individuals who are exposed occupationally, consuming alcohol, smoking and on medication were excluded. The study was approved by the Institutional Ethics Committee and guidelines of Indian Council of Medical Research (ICMR) were followed. The collected blood samples were kept stand alone to separate the plasma for extractions of circulating cell free DNA (ccfDNA) and circulating miRNA (ccfmiRNA). The extracted ccfDNA was further used to evaluate the global DNA methylation levels [5-methylcytosine (5-mC)]. In addition, to isolate lymphocytes the blood was diluted with 0.9 % saline (equal volume of plasma removed), mixed and over layered on Hisep followed by centrifugation at 400g for 30 min (Bhargava et al., 2016). Buffy coat containing lymphocytes was separated into new tube, centrifuged and washed and was further used to extract DNA (for LINE1 analysis), RNA (for analysis of epigenetic modifiers and target genes) and total histone protein (for H3 and H4 analysis) (Gupta et al., 2019). All plasma and extracted samples were stored at −80 °C for further use.

### 2.2. Assessment of global DNA methylation

Cell free circulating DNA (ccfDNA) was extracted from the stand-alone plasma samples by using NucleoSpin® Plasma XS kit. The 100ng of extracted ccfDNA was then evaluated the levels of 5-methylcytosine (5-mC) levels by using MethylFlash™ Global DNA Methylation (5-mC) ELISA Easy Kit following all necessary instructions from the supplier. The readings were recorded using Spark® multimode microplate reader (TECAN, Männedorf, Switzerland) at 450 nm (Mishra et al., 2014). While, methyl specific PCR (MSP) using bisulfite converted DNA was done to check the global methylation status at LINE-1 sequences. The obtained amplicons were then analysed by using agarose gel electrophoresis and the images were captured using UV BioRad Gel Doc™ XR+ (Hercules, California, USA) using Image lab software (Mishra et al., 2010).

### 2.3. Relative gene expression analysis of epigenetic modifiers

The expression levels of epigenetic modifiers DNMT1, DNMT3a, EZH2, EHMT2 or G9a; and p300 was performed by a conventional PCR based approach (Insta Q96™, HiMedia Laboratories, Mumbai, MH, India). Initially, RNA was isolated, quantified and subjected to the cDNA synthesis. The generated cDNA was mixed with NEB Taq 2X master mix and specific primers for amplification on a thermal cycler (GeneAmp® PCR System 9700, Applied Biosystems, USA). After amplification, agarose gel electrophoresis of the amplified products was performed for the analysis and the images were captured using UV BioRad Gel Doc™ XR+ (Hercules, California, USA) using Image lab software (Bhargava et al., 2018b).

### 2.4. Evaluation of H3/H4 histone modifications

Total 21 H3 and 10 H4 modifications were studied by using EpiQuik™ Histone H3/H4 Modification Multiplex Assay Kits. The studied H3 modifications include mono-, di-, and trimethylation of lysine residues at 4, 9, 27, 36, and 79; mono-, di-, and tri-acetylation of lysine residues at 9, 14, 18, and 56 position; and phosphorylation of serine residue at 10 and 28. While, studied H4 modifications includes mono-, di- and tri-methylation of lysine 20; acetylation of lysine 5, 8, 12 and 16; phosphorylation of serine 1; symmetric and asymmetric di-methylation of arginine 3. The readings were recorded at 450 nm on Spark® multimode microplate reader (TECAN, Männedorf, Switzerland) (Bhargava et al., 2019).

### 2.5. miRNA expression profiling

The expression profiling of a selected miRNA panel, which include Let-7a, Let-7b-5p, Let-7d, Let-7e, miR-16-5p, miR-17, miR-27, miR-28-5p, miR-29a, miR-98, miR-128-2, miR-142-5p, miR-200c, miR-202, miR-221 and miR-451a was performed using the method described earlier. In brief, isolated miRNA was subjected to poly-adenylation, cDNA synthesis and amplification by using respective primers on an Insta Q-96™ real-time PCR (HiMedia Laboratories, Mumbai, MH, India). We used U6 as an internal control. After completion of the programmed PCR cycles, Ct values were recorded and dCt values were obtained by subtracting the Ct value of internal control from the Ct value of the miRNAs (Shandilya et al., 2020).

### 2.6. Analysis of miRNA targets

A panel of 85 genes including AKT3, CDK2, CDK4, CDK6, CDKN1B, CDKN2A, CDKN2B, LAMC3, PIAS3, RASSF1, CHUK, ITGA6, COL4A2, E2F1, E2F2, E2F3, EGF, ERBB2, AKT1, AKT2, FHIT, FOXO3, FN1, PIK3R5, LAMA1, GRB2, APAF1, BIRC2, BIRC3, XIAP, IKBKB, ARAF, LAMA2, LAMB2, LAMC2,MAX, NFKBIA, NOS2, NRAS, PIAS4, PDPK1, PIK3CA, PIK3R1, PLCG1, PLCG2, CYCS, PRKCA, PRKCB, PRKCG, MAPK1, MAP2K1, BAD, PTEN, PTGS2, PTK2, RAF1, RARB, RB1, CCND1, BCL2, RELA, BCL2L1, RXRA, RXRB, RXRG, SKP2, SOS1, SOS2, BRAF, STK4, TGFA, TP53, TRAF1, ITGAV, RASSF5, CASP9, PIK3R3, IKBKG, PIAS1, CCNE1, PIAS2, CCNE2, TRAF4, and HRAS was selected as miRNA targets. While, GUSB glucuronidase beta was used as internal control. The analysis was performed by using KAPA SYBR FAST One-Step qPCR kit on an Insta Q-96™ real-time PCR (HiMedia Laboratories, Mumbai, MH, India). After PCR cycles, dCt values were obtained by subtracting Ct value of internal control to the Ct value of the test gene. ddCT values were obtained by reducing the dCt (Low-risk) from dCt (high-risk) and fold change was calculated by using formula 2^A(dclct)^(Bhargava et al., 2020).

### 2.11. Statistical analysis

Data was analysed using GraphPad Prism 8. Significance was calculated by employing unpaired t test with Welch’s correction and two-way ANOVA with multiple comparison tests.

## 3. Results

### 3.1. Global DNA methylation

DNA methylation is a vital epigenetic process that regulates the functioning vital genes of cellular machinery. The higher gene methylation reduces accessibility of transcription machinery at the transcription site and causes gene silencing, while the hypomethylation leads to gene activation. Upon quantitative analysis, the observed mean levels of % 5-mC in the individuals living at high-risk regions were 22.005 ± 1.041 % and for low-risk regions, the levels were 16.769 ± 0.890 % (Figure 1). In addition, as compared to low-risk population, the significantly higher levels of LINE-1 methylation, observed among the high-risk population further confirmed the above results (Data not shown). Combined together, our results suggested a hypermethylated DNA profile in the high-risk group as compared to the low-risk group.

**Figure 1:**
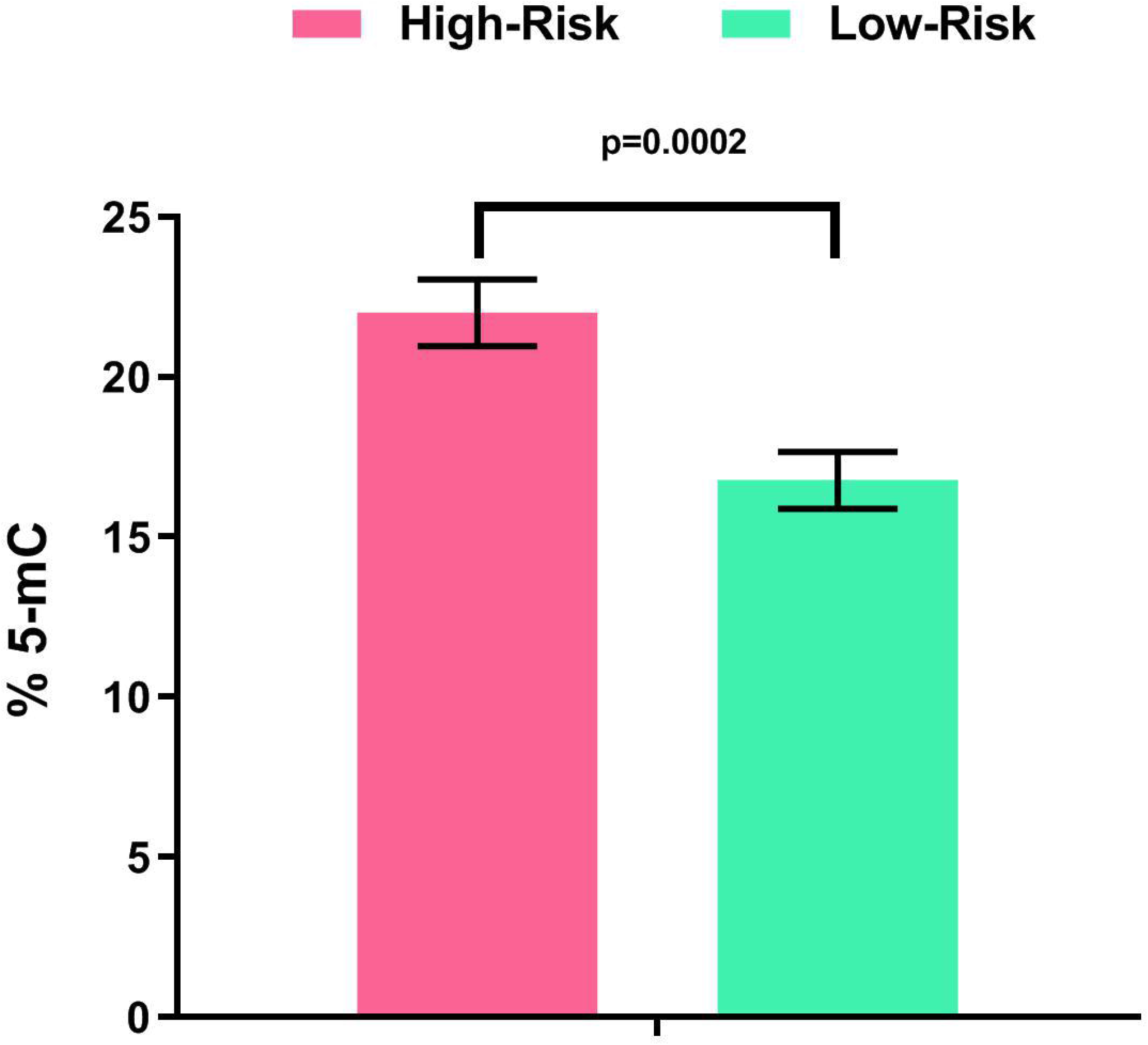
DNA methylation levels. Graph showing global DNA methylation levels assessed through percent 5-mC in circulating DNA of the individuals residing at high-risk and low-risk regions. Data expressed as mean ± SE and p ≤ 0.05 was considered statistically significant.

### 3.2. Status of epigenetic modifiers

Epigenetic modifiers such as DNMTs, histone acetyltransferase and methyltransferase are the crucial regulators of different epigenetic modifications. For instance, DNMTs catalyse the covalent transfer of a methyl group to the C-5 position of the cytosine ring to maintain the DNA methylation levels, while, histone acetyltransferase catalytically adds the acetyl group to the lysine residue histones. Similarly, the histone methyl transferase, functions to add methyl group to the lysine and arginine residues of histone proteins. In comparison to controls, we observed higher expression of different epigenetic modifiers i.e. DNMT1, DNMT3a, EZH2, p300 and G9a among the high-risk group. The observed band intensities were comparatively higher in the high-risk group than those of low risk group (Figure 2). As these epigenetic modifiers precisely regulates different epigenetic processes including DNA methylation and covalent histone modifications, such dysregulation of epigenetic modifiers might result in aberrant epigenetic landscapes and transcriptional programs.

**Figure 2:**
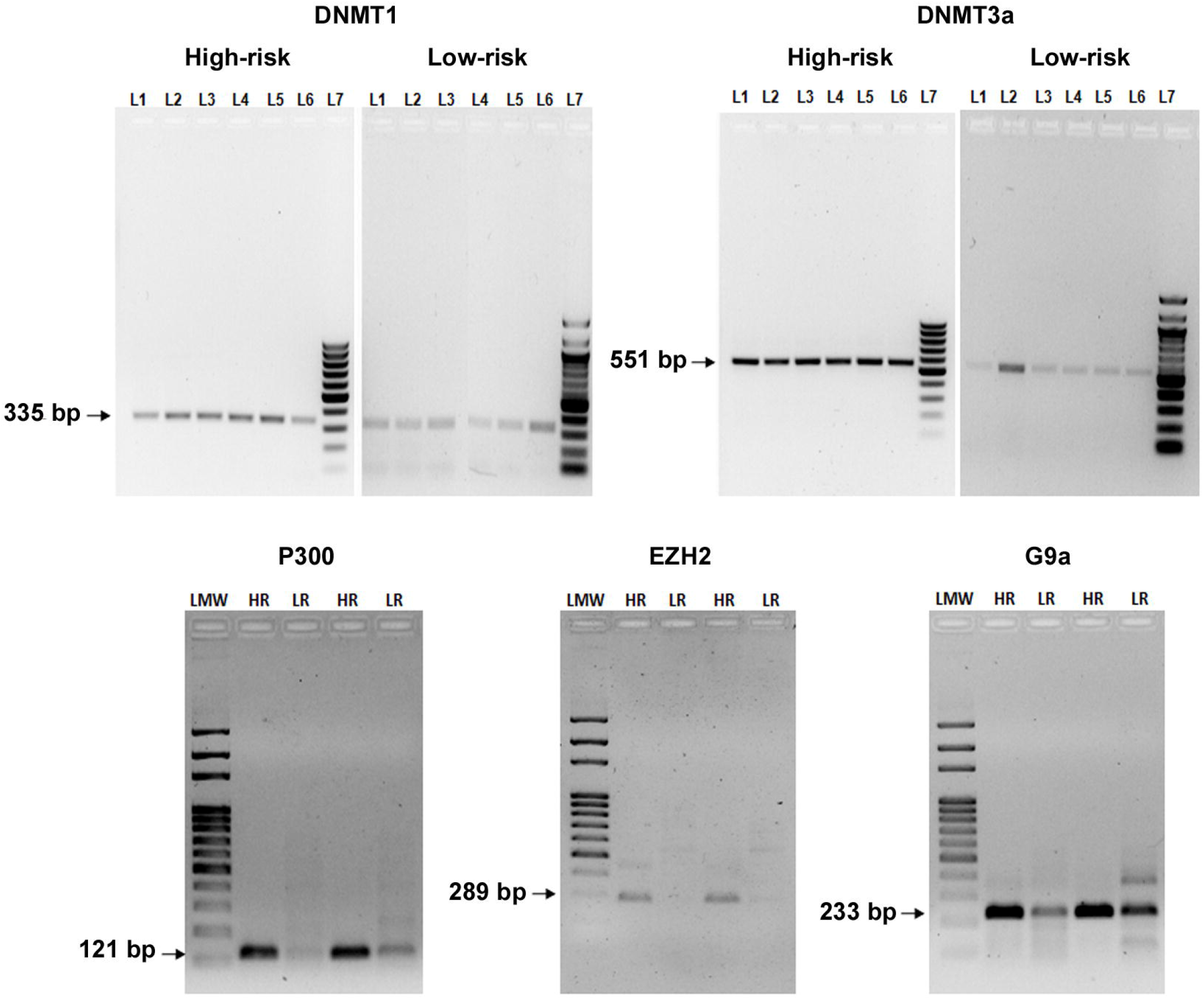
Epigenetic modifiers. The representative gel images showing DNMT1, DNMT3a, P300, EZH2 and G9a expression patterns in peripheral lymphocytes of individuals residing at high-risk and low-risk regions. L1 (Lane) 1 - L6: Samples, L7: Low molecular weight marker, HR: high-risk, LR: low-risk, LMW: low molecular weight marker.

### 3.3. Aberrant histone modification patterns

Histone modifications are the major regulatory processes at the epigenomic level, which help to orient chromatin in different configurations depending upon the modification of amino residue of histone and forms the basis of histone codes. Histone 3 and 4 play important role in the regulation of gene transcription and are closely linked with the process of DNA methylation. We observed aberrant levels of H3 and H4 modifications in the individuals residing at high-risk regions. In comparison to low risk group, a significant increase in the levels of H3K4me2 and H3K9ac was found in the high-risk group (Figure 3a). While, a non-significant increase in the levels of all 10 H4 modifications was observed in the high-risk group individuals, when compared with the low risk group (Figure 3b).

**Figure 3:**
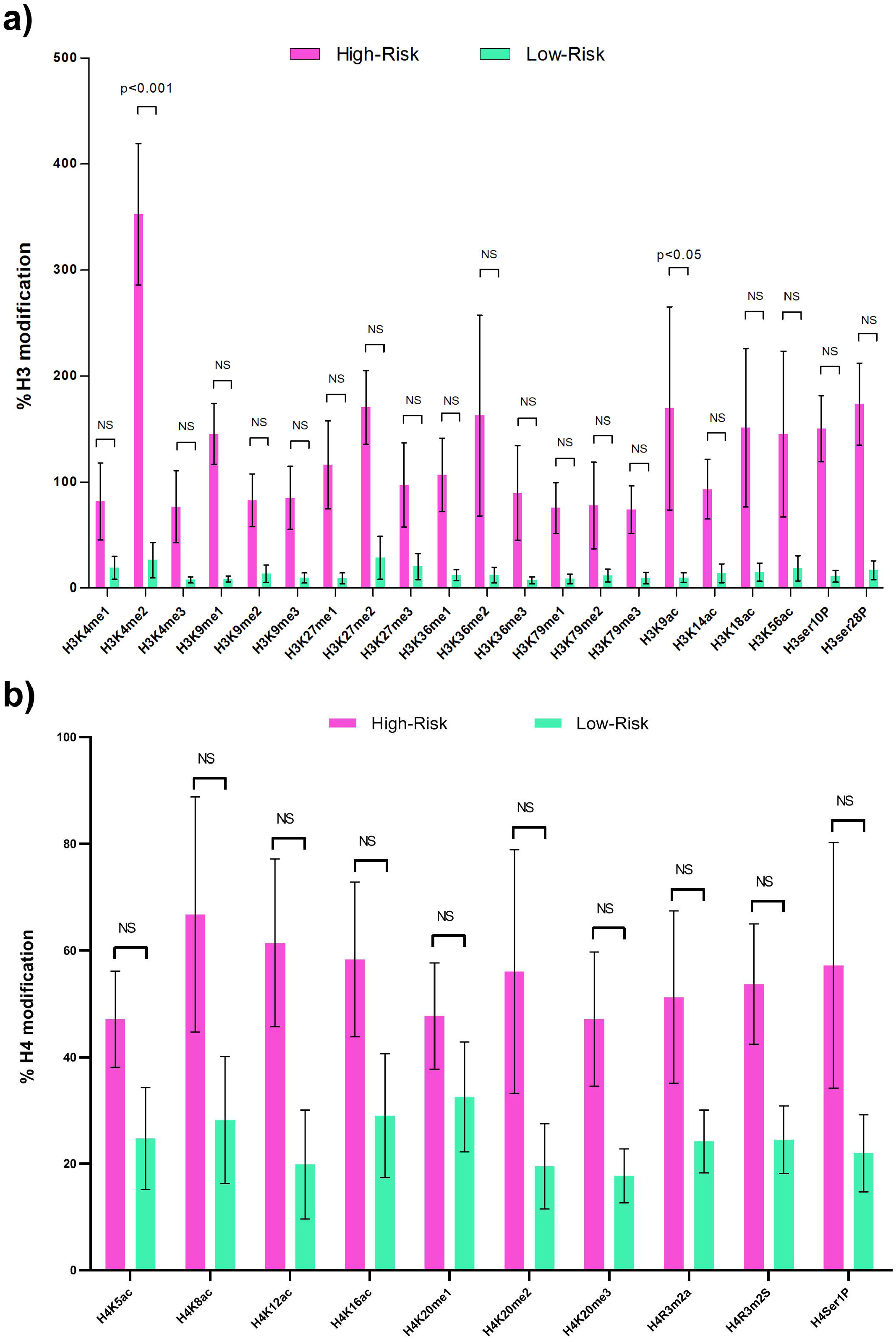
Histone (H3 and H4) modifications. Graph showing the histone H3 and H4 modifications in the peripheral lymphocytes of the individuals residing in high-risk and low-risk regions. Data expressed as mean ± SE and p < 0.05 was considered significant as compared with controls.

### 3.4. Differential miRNA expression profile

Interconnections of miRNAs to the other epigenetic processes such as DNA methylation and histone modification, signifies their mechanistic role as epigenetic regulators. The quantitative analysis of the expression of 16 circulating miRNAs showed marked alterations among high-risk group. The results showed that in comparison to low risk group the expression of let-7a, let-7b, miR-17, miR-28, miR-98, and miR-200c in high risk group were up-regulated while the levels of let-7d, let-7e, miR-16-5p, miR-128, miR-145-5p, and miR-202 were observed downregulated (Figure 4).

**Figure 4:**
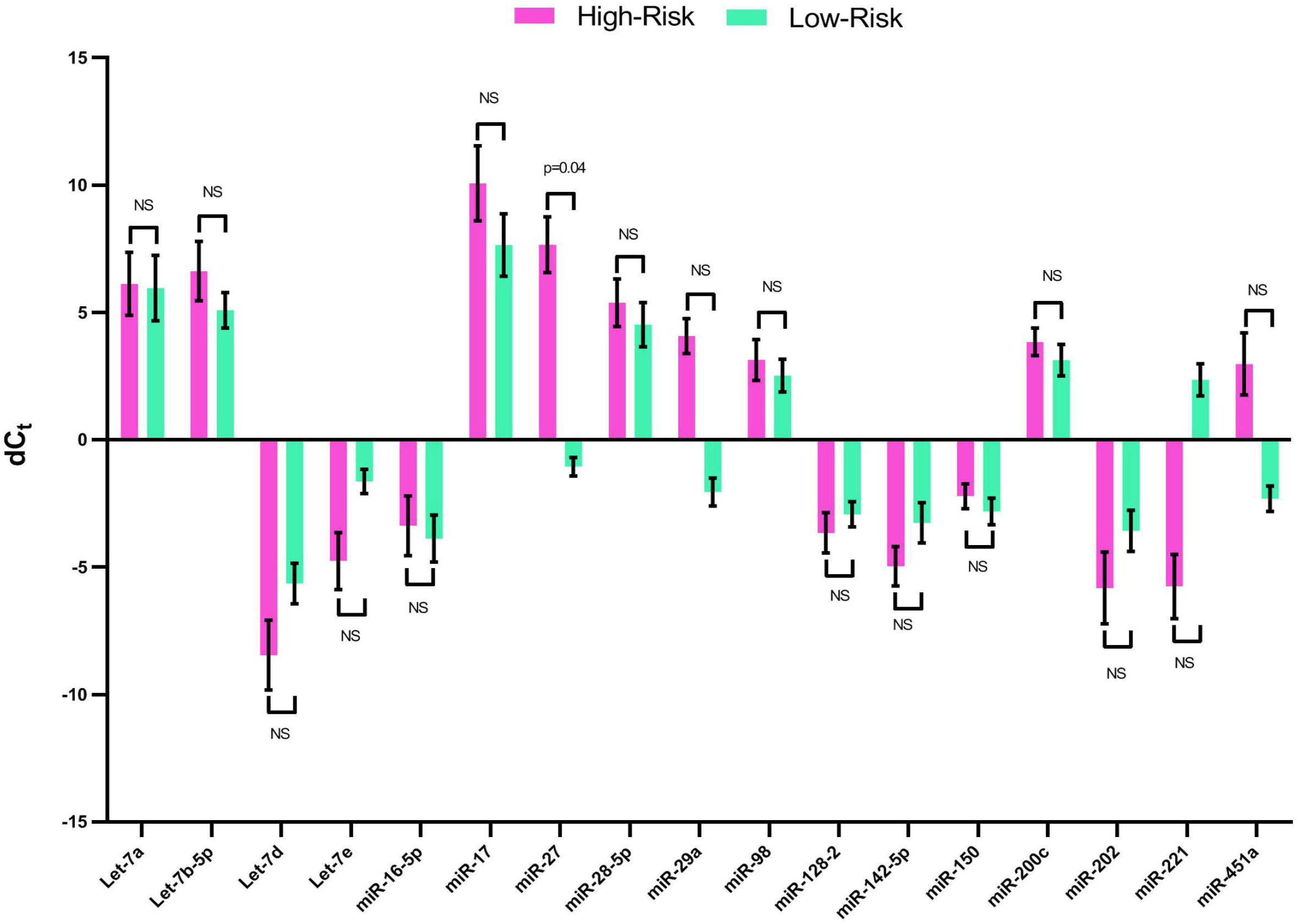
miRNA profiling. Graph showing circulating miRNA profile in individuals residing at high-risk and low-risk regions of India. Data expressed as mean ± SE and p < 0.05 was considered significant. dCt values were obtained by subtracting the Ct value of internal control from the Ct value of the miRNAs

### 3.5. Target gene analysis

Primarily function of the miRNA involves complementary based inhibition or degradation of the mRNA thereby limits the translation of the mRNA. Although, these regulated processes require and work in coordinated way with the several proteins that may be the transcription factor, intermediate proteins, accessory proteins, messenger proteins and any other protein connecting cascade reactions regulating certain cellular processes. The analysis of the genes (85) involved in the vital signalling pathways revealed marked alteration in the levels of expression of CDK2/4, CDKN1B/2B, RASSF1, ITGA6, COL4A2, FHIT, FOXO3, BIRC2/3, NRAS, PIK3R1, PLCG1, BCL2, CCND1, SKP2 and ITGAV gene transcripts in high risk group when compared with the low risk group (Figure 5). Importantly, these gene transcripts are known to functionally regulate cell cycle, inflammation, cell survival, apoptosis and cell adhesion.

**Figure 5:**
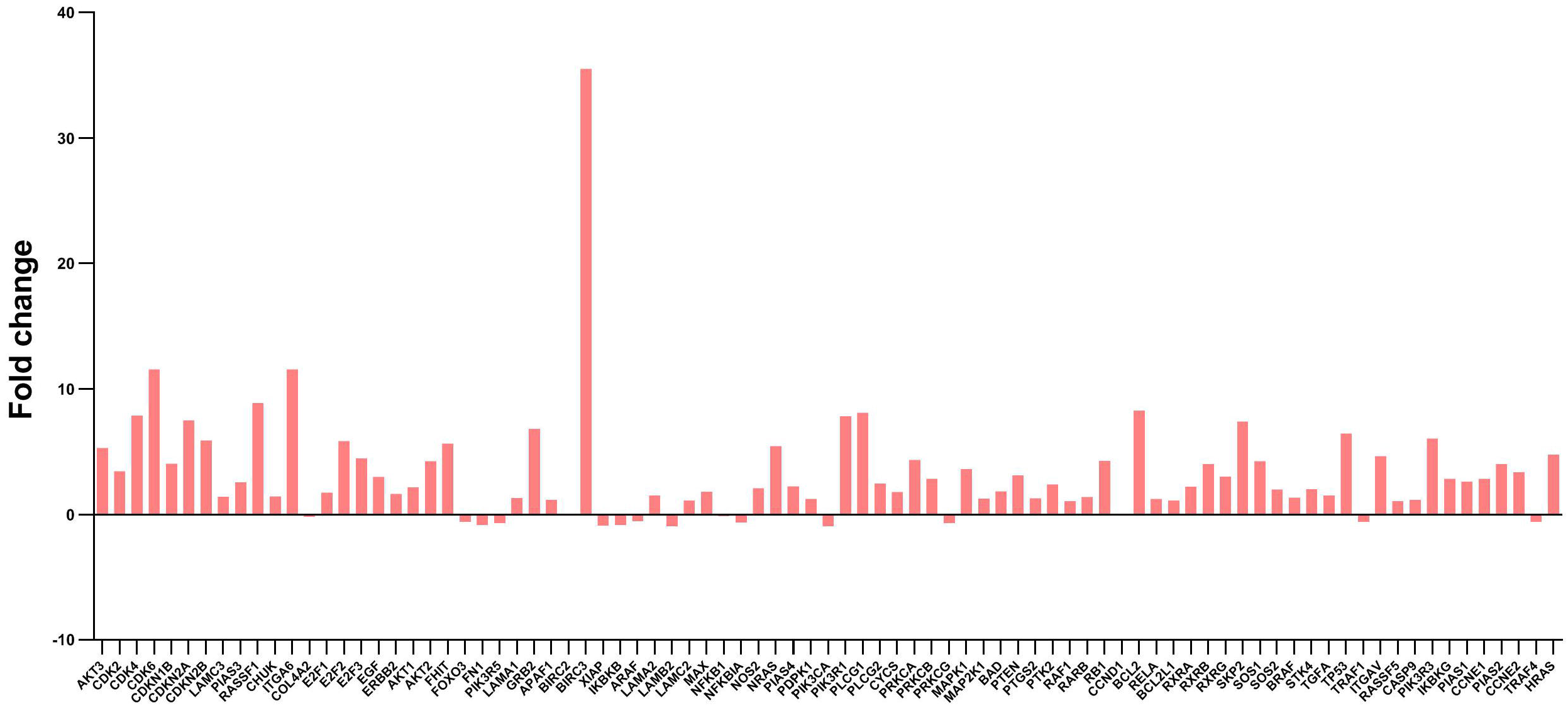
Target gene analysis. Graph showing the fold change in the expression of miRNA target genes in the peripheral lymphocytes of the individuals residing at high-risk and low-risk regions. The fold change was calculated as 2^^ΔΔCT^ by identifying (dC_T_) as the difference of internal control from their respective control and test values.

## 4. Discussion

Environmental stress can potentially modify the pre-existing genetic programs, leading to a number of biological or adverse health outcomes. Compelling evidences including earlier work from our laboratory suggest that exposure to environmental pollutants such as air-borne PM disturbs mitochondrial redox signaling and induces a PI3 kinase mediated DNA damage response (Bhargava et al., 2018b). It further stimulates an mtDNA-ROS-NF-KP pathway to cause alterations within the pattern of epigenetic markers that eventually regulate the chromatin structure or gene expression (Bhargava et al., 2019). Such causalities in the vital epigenetic programs may interrupt distinct cellular activities, biological functions and cell-fate decisions, resulting into aberrant phenotypes. To shed light on the possible impacts of air-borne PM and to find out important clues relating to exposure, we herein characterized epigenomic signatures in the individuals living in high and low-risk air pollution zones. Collectively, our results provide first insights of aberrant epigenomic signatures in peripheral circulation of the individuals residing in the high-risk air pollution zones.

Epigenetic modifications that primarily involves DNA methylation, post translational histone modifications, and noncoding RNAs, stands at the interface of the human genome, development and environmental exposure (Bunkar et al., 2016). These processes often mediate gene-environment interactions and can alter the ongoing cellular mechanisms to facilitate the adaptation of a cell to its environment. Interestingly, epigenetic modifications may remain preserved among the circulating nucleic acids aroused primarily from cell death and can be used to generate an epigenome-wide analytical landscape comprising essential insights of an individual’s disease status (Bhargava et al., 2017; Bhargava et al., 2018a). It is now evident that the exposure to different environmental pollutants, such as PM and residual organic compounds can cause epigenetic imbalances (Shukla et al., 2019; Bhargava et al., 2020). However, during such processes, the components of pollutants, which have varying extents of health impacts and toxicity, have an important role. In particular, the metal components of PM like Nickel, Arsenic, Lead, Cobalt and Chromium act as pro-oxidant and induce generation of ROS and inflammatory responses. This metallic component significantly disrupts mitochondrial machinery, alters the functioning of respiratory chain complexes and induces the premature release of electrons. This results in the disruption of proton motive force, shift in metabolic concentration and induction of DNA damage response pathways (Bhargava et al., 2018b). In a developing country like India, where PM concentration has reached unprecedented levels, close association of metallic components with PM is a matter of significant concern. The WHO has categorized Delhi, Gwalior & Raipur among the highly polluted cities of India and PM in these cities were found to remain associated with organic carbon, inorganic ionic components and metals including lead, chromium, cadmium, and nickel (Hazarika et al., 2015; Jain et al., 2020).

Upon induction of DNA damage response pathways, the mobilization of damage repair proteins further catalyses the recruitment of DNMTs to the damage site, which may result in the attachment of the methyl groups at the 50-carbon of cytosine residues located in the CG dinucleotides (Shukla et al., 2019). Recent studies have also shown that exposure to PM causes alterations in the DNA methylation profile of the exposed cells (Bhargava et al., 2019; Lee et al., 2019). These changes are mainly associated with the variations in mitochondrial metabolite pools, which are likely to affect the functioning of enzymes regulating the epigenetic landscape. Importantly, DNA methylation is one of the best-understood epigenetic features in case of PM-associated diseases. Accumulating evidences have suggested the link of altered DNA methylation patterns with the prevalence of respiratory diseases cardiovascular disorders and cancer (Yan et al., 2020; Tessema et al., 2020; Liu et al., 2020). In addition, the detection of methylated DNA circulating in the peripheral blood has gained significant attention for the cancer diagnosis, specifically in the determination of early, late as well as stage of the malignancy (Li et al., 2019b). Therefore, understanding such mechanisms in circulation may prove vital to establish a possible link between long-term ambient PM exposures and associated non-communicable diseases. In our study, the observed levels of global DNA methylation among the individuals residing at the high-risk regions suggested a hyper-methylation profile. These results strongly coincided with the expression levels of DNA methylation enzymes (Figure 1, 2).

Mechanistically, the process of DNA methylation is closely associated with the post-translational histone modifications that involve methylation, acetylation, ubiquitination, phosphorylation, and/or sumoylation. Specific epigenetic modifiers catalyze all these modifications in histones tails during replication and transcription to increase the accessibility of the DNA (Jeltsch et al., 2018). However, depending upon the type of the modification, it may result in either gene activation or repression to regulate vital cellular processes such as cellular differentiation, cell cycle progression, DNA replication and apoptosis (Ding et al., 2016; Lee et al., 2019). An imbalance in the regulation of these mechanisms can disturb the equilibrium, which in turn may result in generation of specific disease associated proteomes. Studies suggest that exposure to PM and its components can alter a number of such processes including increase in the methylation (di and tri) and acetylation mark on the histone H3 and H4 (Liu et al., 2015; Kresovich et al., 2017). Both these (H3 and H4) modifications are important, as they are considered as biomarkers of different diseases including cancer. While, the status of epigenetic modifiers regulating histone modifications following exposure to air-borne PM is not well explored, it is assumed that hypermethylation of CpG sites located at gene promoters are commonly associated with transcriptional silencing. This is mainly through influencing the binding of transcription factors to their targets and increasing the affinity to methylated DNA binding proteins that assist the further recruitment of other epigenetic modifiers. In light of this, we assessed the levels of histone modifiers in the present study and observed their enhanced levels in the high-risk population (Figure 2). Upon further assessment of histone modifications, the results suggested the occurrence of significantly higher levels of H3K4me2 and H3K9ac, in the high-risk population when compared to the low risk (Figure 3). The observations are of great importance, as these modifications can alter the transcriptional status by synergistically influencing accessibility of transcription factors or RNA polymerase II and chromatin structure.

Moreover, in recent times, miRNAs, which regulate mRNA silencing and posttranscriptional gene expression, has emerged as one of the important epigenetic molecules. These small noncoding RNA molecules remains associated with other epigenetic events and functions as main epigenetic regulator of various crucial cellular events (Morales et al., 2017). Importantly, the crosstalk between DNA methylation and miRNAs acts as an essential epigenetic characteristic that can serve as a bridge between the air-pollutant exposure and associated health outcomes. On the one hand, hypo-methylation or hypermethylation causes activation/repression of onco-miRs/tumor-suppressor miRNAs; while on the other hand miRNAs can target DNMTs or methylation-related proteins and regulate DNA methylation thus reflecting the potential of miRNAs to regulate disease development. Notably, few studies also suggest a linear relationship of altered miRNA expression patterns with air pollutant exposure (Alfano et al. 2018; Espín-Pérez et al., 2018; Xiao et al., 2019). It is assumed that PM induced oxidative stress may probably alter the pri-miRNA structure, inhibit the function of Dicer and act as intermediate molecule to activate the stress-related transcription factors such as p53, NF-κβ, FOXO and HIF. These factors may further transactivate downstream miRNAs (Jaksik et al., 2014, Hoffend et al., 2017). Owing to the role of aberrantly expressed miRNAs in the onset of different PM associated diseases, we herein assessed the expression profile 17 miRNAs important to regulate vital cellular mechanisms. The results observed herein were in agreement with the above observations, that exposure to air-borne PM alters the expression of miRNAs. In comparison to low risk group, the high-risk group showed higher degree of changes in expression profile of all studied miRNAs (Figure 4). Such alterations in PM exposed subjects may trigger a cascade of events that are closely associated with the disturbances in various complex biological processes including inflammation, endothelial dysfunction, and coagulation (Fossati et al., 2014). However, there is far less understanding about the miRNA associated complex molecular networks in individuals exposed to PM. In our study, the possibilities of deregulation in the vital signalling mechanisms was confirmed by the analysis of 85 genes panel, which revealed differential expression of the gene transcripts regulating cell cycle, inflammation, cell survival, apoptosis and cell adhesion in the individuals living in high-risk zones (Figure 5).

In conclusion, our results provided first evidence for explaining the air-borne PM associated epigenetic mechanisms using a multi-city approach in India. We observed aberrant levels of epigenetic modifications in peripheral circulation of the individuals living in high-risk zones of the country. As epigenetic mechanisms control various facets of chromatin structure and plays a functional role in transcriptional regulation of vital genes, it is evident that improved understanding of these signatures in circulation will provide information about ongoing cellular changes. It also forms the background of using these molecules as a biomarker for risk assessment of air-borne PM associated diseases. Interestingly, there is an increasing probability that such modifications in the epigenome may remain coded in a way that the information can be transferred to the subsequent generations. However, unavailability of a proved mechanism certainly questions the occurrence of such complicated phenomenon. Future studies with larger sample size are highly warranted to confirm the current findings and identify specific pathways affected by exposure-related changes in circulating epigenetic signatures, as well as to link these observations with air pollution-associated disease outcomes. Further steps to develop a point-of-care epigenomic assay for human bio-monitoring may be immensely beneficial to reduce the health burden of air pollution especially in lower-middle-income countries such as India.

**Table 1:** Demographic data of individuals recruited from the high and low air pollution areas.

| **Variables** | **High-Risk** | **Low-Risk** |
| --- | --- | --- |
| **Number** | 60 | 60 |
| **Age (Mean±SD)** | 34.88±<br>15.16 yrs | 35.45±<br>16.77 yrs |
| **Years of residence** | 17.58±<br>3.52 yrs | 18.26±<br>2.89 yrs |
| **Marital status** |  |  |
| **Married** | 41 | 40 |
| **Unmarried** | 19 | 20 |
| **Literacy status** |  |  |
| **Basic** | 32 | 41 |
| **Graduate** | 19 | 15 |
| **Post-graduate** | 9 | 3 |
| **Occupation** |  |  |
| **Housewife** | 29 | 28 |
| **Student** | 15 | 16 |
| **Job** | 7 | 12 |
| **Working hours** | ≥8 hrs | ≥5 hrs |
| **Duration** | ≥6 yrs | ≥ 12 yrs |
| **Business** | 9 | 4 |
| **Working hours** | ≥12 | ≥12 |
| **Duration** | ≥12 | ≥12 |
| **Family income** |  |  |
| **≥20000** | 5 | 16 |
| **≥30000** | 20 | 21 |
| **≥50000** | 35 | 23 |
| **Type of exposure** |  |  |
| **Indoor** | 13 | 31 |
| **Outdoor** | 47 | 29 |
| **Occupational exposure** | None | None |
| **Fuel type used** |  |  |
| **Non-solid-electric/LPG** | 60 | 60 |
| **Kerosene oil/ Solid fuels** | None | None |
| **Have separate kitchen** | Yes | Yes |
| **House type** |  |  |
| **Concrete** | Yes | Yes |
| **Mud/wood** | None | None |

## Data Availability

Yes

## Authors Contributions

PKM devised the concept, developed the methodology and supervised the experiments; NB and RDS performed the DNA methylation assays; NB and RK carried out the miRNA profiling and target-gene expression analysis; PKG, LL and AB carried out histone characterization; PKM, NB, LL and PKG performed demographic analysis; PKM, RT and KC analyzed and interpreted the data; and PKM, NB and AB drafted the manuscript.

## Acknowledgements

PKM and KC are thankful to the Ministry of Health & Family Welfare (MoHFW) and Ministry of Human Resource & Development (MHRD), Government of India, New Delhi for funding support received through IMPRINT India Initiative (Project ID-4352).

## References

1. Alfano R, Herceg Z, Nawrot TS, Chadeau-Hyam M, Ghantous A, Plusquin M. The Impact of Air pollution on our epigenome: How far is the evidence? (A systematic review). Curr Environ Health Rep. 2018; 5: 544–578. doi: 10.1007/s40572-018-0218-8.

2. Bhargava A, Shukla A, Bunkar N, Shandilya R, Lodhi L, Kumari R, Gupta PK, Rahman A, Chaudhury K, Tiwari R, Goryacheva IY, Mishra PK. Exposure to ultrafine particulate matter induces NF-κβ mediated epigenetic modifications. Environ Pollut. 2019; 252(Pt A): 39-50. doi: 10.1016/j.envpoL2019.05.065.

3. Bhargava A, Bunkar N, Aglawe A, Pandey KC, Tiwari R, Chaudhury K, Goryacheva IY, Mishra PK. Epigenetic Biomarkers for risk assessment of particulate matter associated lung cancer. Curr Drug Targets. 2018a; 19: 1127-1147. doi: 10.2174/1389450118666170911114342.

4. Bhargava A, Khare NK, Bunkar N, Chaudhury K, Pandey KC, Jain SK, Mishra PK. Cell-Free circulating epigenomic signatures: non-invasive biomarker for cardiovascular and other age-related chronic diseases. Curr Pharm Des. 2017; 23: 1175–1187. doi: 10.2174/1381612822666161027145359.

5. Bhargava A, Khare NK, Bunkar N, Lenka RK, Mishra PK. Role of mitochondrial oxidative stress on lymphocyte homeostasis in patients diagnosed with extra-pulmonary tuberculosis. Cell Biol Int. 2016; 40: 166–76. doi: 10.1002/cbin.10549.

6. Bhargava A, Kumari R, Khare S, Shandilya R, Gupta PK, Tiwari R, Rahman A, Chaudhury K, Goryacheva IY, Mishra PK. Mapping the mitochondrial regulation of epigenetic modifications in association with carcinogenic and noncarcinogenic polycyclic aromatic hydrocarbon exposure. Int J Toxicol. 2020: 1091581820932875. doi: 10.1177/1091581820932875. PMID: 32588678

7. Bhargava A, Tamrakar S, Aglawe A, Lad H, Srivastava RK, Mishra DK, Tiwari R, Chaudhury K, Goryacheva IY, Mishra PK. Ultrafine particulate matter impairs mitochondrial redox homeostasis and activates phosphatidylinositol 3-kinase mediated DNA damage responses in lymphocytes. Environ Pollut. 2018b; 234: 406-419. doi: 10.1016/j.envpol.2017.11.093.

8. Bunkar N, Pathak N, Lohiya NK, Mishra PK. Epigenetics: A key paradigm in reproductive health. Clin Exp Reprod Med. 2016; 43: 59–81. doi: 10.5653/cerm.2016.43.2.59.

9. Chen X, Guo J, Huang Y, Liu S, Huang Y, Zhang Z, Zhang F, Lu Z, Li F, Zheng JC, Ding W. Urban airborne PM_2_._5_-activated microglia mediate neurotoxicity through glutaminase-containing extracellular vesicles in olfactory bulb. Environ Pollut. 2020; 264: 114716. doi: 10.1016/j.envpol.2020.114716.

10. Chen Y, Shen H, Smith KR, Guan D, Chen Y, Shen G, Liu J, Cheng H, Zeng EY, Tao S. Estimating household air pollution exposures and health impacts from space heating in rural China. Environ Int. 2018; 119:117-124. doi: 10.1016/j.envint.2018.04.054.

11. Cheng M, Wang B, Yang M, Ma J, Ye Z, Xie L, Zhou M, Chen W. microRNAs expression in relation to particulate matter exposure: A systematic review. Environ Pollut. 2020; 260: 113961. doi: 10.1016/j.envpoL2020.113961.

12. Chu C, Zhang H, Cui S, Han B, Zhou L, Zhang N, Su X, Niu Y, Chen W, Chen R, Zhang R, Zheng Y. Ambient PM2.5 caused depressive-like responses through Nrf2/NLRP3 signaling pathway modulating inflammation. J Hazard Mater. 2019; 369:180-190. doi: 10.1016/j.jhazmat.2019.02.026.

13. Ding R, Jin Y, Liu X, Zhu Z, Zhang Y, Wang T, Xu Y. H3K9 acetylation change patterns in rats after exposure to traffic-related air pollution. Environ Toxicol Pharmacol. 2016; 42:170-5.

14. Duan H, Jia X, Zhai Q, Ma L, Wang S, Huang C, Wang H, Niu Y, Li X, Dai Y, Yu S, Gao W, Chen W, Zheng Y. Long-term exposure to diesel engine exhaust induces primary DNA damage: a population-based study. Occup Environ Med. 2016; 73: 83–90. doi: 10.1136/oemed-2015-102919.

15. Espín-Pérez A, Krauskopf J, Chadeau-Hyam M, van Veldhoven K, Chung F, Cullinan P, Piepers J, van Herwijnen M, Kubesch N, Carrasco-Turigas G, Nieuwenhuijsen M, Vineis P, Kleinjans JCS, de Kok TMCM. Short-term transcriptome and microRNAs responses to exposure to different air pollutants in two population studies. Environ Pollut. 2018; 242(Pt A): 182-190. doi: 10.1016/j.envpol.2018.06.051.

16. Eze IC, Jeong A, Schaffner E, Rezwan FI, Ghantous A, Foraster M, Vienneau D, Kronenberg F, Herceg Z, Vineis P, Brink M, Wunderli JM, Schindler C, Cajochen C, Röösli M, Holloway JW, Imboden M, Probst-Hensch N. Genome-Wide DNA Methylation in Peripheral Blood and Long-Term Exposure to Source-Specific Transportation Noise and Air Pollution: The SAPALDIA Study. Environ Health Perspect. 2020; 128: 67003. doi: 10.1289/EHP6174.

17. Feinberg AP. The Key Role of Epigenetics in Human Disease Prevention and Mitigation. N Engl J Med. 2018; 378: 1323–1334. doi: 10.1056/NEJMral402513.

18. Fossati S, Baccarelli A, Zanobetti A, Hoxha M, Vokonas PS, Wright RO, Schwartz J. Ambient particulate air pollution and microRNAs in elderly men. Epidemiology. 2014; 25: 68–78. doi: 10.1097/EDE.0000000000000026.

19. Gupta P, Bhargava A, Kumari R, Lodhi L, Tiwari R, Gupta PK, Bunkar N, Samarth R, Mishra PK. Impairment of Mitochondrial-Nuclear Cross Talk in Lymphocytes Exposed to Landfill Leachate. Environ Health Insights. 2019; 13: 1178630219839013. doi: 10.1177/1178630219839013.

20. Hazarika N, Jain VK, Srivastava A. Source identification and metallic profiles of size-segregated particulate matters at various sites in Delhi. Environ Monit Assess. 2015; 187: 602. doi:10.1007/s10661-015-4809-7

21. Hoffend NC, Magner WJ, Tomasi TB. The epigenetic regulation of Dicer and microRNA biogenesis by Panobinostat. Epigenetics. 2017; 12: 105–112. doi: 10.1080/15592294.2016.1267886.

22. India State-Level Disease Burden Initiative Air Pollution Collaborators. The impact of air pollution on deaths, disease burden, and life expectancy across the states of India: the Global Burden of Disease Study 2017. Lancet Planet Health. 2019; 3: e26-e39. doi:10.1016/S2542-5196(18)30261-4.

23. Jain S, Sharma SK, Vijayan N, Mandal TK. Seasonal characteristics of aerosols (PM2.5 and PM10) and their source apportionment using PMF: A four year study over Delhi, India. Environ Pollut. 2020; 262: 114337. doi:10.1016/j.envpol.2020.114337.

24. Jaksik R, Lalik A, Skonieczna M, Cieslar-Pobuda A, Student S, Rzeszowska-Wolny J. MicroRNAs and reactive oxygen species: are they in the same regulatory circuit? Mutat Res Genet Toxicol Environ Mutagen. 2014; 764-765: 64-71. doi: 10.1016/j.mrgentox.2013.09.003.

25. Jeltsch A, Broche J, Bashtrykov P. Molecular processes connecting DNA methylation patterns with DNA methyltransferases and histone modifications in mammalian genomes. Genes (Basel). 2018; 9:566. doi: 10.3390/genes9110566.

26. Kresovich JK, Zhang Z, Fang F, Zheng Y, Sanchez-Guerra M, Joyce BT, Zhong J, Chervona Y, Wang S, Chang D, McCracken JP, Díaz A, Bonzini M, Carugno M, Koutrakis P, Kang CM, Bian S, Gao T, Byun HM, Schwartz J, Baccarelli AA, Hou L. Histone 3 modifications and blood pressure in the Beijing Truck Driver Air Pollution Study. Biomarkers. 2017; 22: 584-593.

27. Landrigan PJ, Fuller R, Acosta NJR, Adeyi O, Arnold R, Basu NN, Baldé AB, Bertollini R, Bose-O’Reilly S, Boufford JI, Breysse PN, Chiles T, Mahidol C, Coll-Seck AM, Cropper ML, Fobil J, Fuster V, Greenstone M, Haines A, Hanrahan D, Hunter D, Khare M, Krupnick A, Lanphear B, Lohani B, Martin K, Mathiasen KV, McTeer MA, Murray CJL, Ndahimananjara JD, Perera F, Potocnik J, Preker AS, Ramesh J, Rockstrbm J, Salinas C, Samson LD, Sandilya K, Sly PD, Smith KR, Steiner A, Stewart RB, Suk WA, van Schayck OCP, Yadama GN, Yumkella K, Zhong M. The Lancet Commission on pollution and health. Lancet. 2018; 391: 462–512. doi: 10.1016/S0140-6736(17)32345-0.

28. Lee MK, Xu CJ, Carnes MU, Nichols CE, Ward JM; Bios consortium, Kwon SO, Kim SY, Kim WJ, London SJ. Genome-wide DNA methylation and long-term ambient air pollution exposure in Korean adults. Clin Epigenetics. 2019; 11: 37.

29. Li D, Zhang R, Cui L, Chu C, Zhang H, Sun H, Luo J, Zhou L, Chen L, Cui J, Chen S, Mai B, Chen S, Yu J, Cai Z, Zhang J, Jiang Y, Aschner M, Chen R, Zheng Y, Chen W. Multiple organ injury in male C57BL/6J mice exposed to ambient particulate matter in a real-ambient PM exposure system in Shijiazhuang, China. Environ Pollut. 2019a; 248: 874-887. doi: 10.1016/j.envpol.2019.02.097.

30. Li J, Xing X, Zhang X, Liang B, He Z, Gao C, Wang S, Wang F, Zhang H, Zeng S, Fan J, Chen L, Zhang Z, Zhang B, Liu C, Wang Q, Lin W, Dong G, Tang H, Chen W, Xiao Y, Li D. Enhanced H3K4me3 modifications are involved in the transactivation of DNA damage responsive genes in workers exposed to low-level benzene. Environ Pollut. 2018; 234: 127–135. doi: 10.1016/j.envpol.2017.11.042.

31. Li L, Fu K, Zhou W, Snyder M. Applying circulating tumor DNA methylation in the diagnosis of lung cancer. Precision Clinical Medicine, 2019b, 1-12. doi: 10.1093/pcmedi/pbz003

32. Liu C, Xu J, Chen Y, Guo X, Zheng Y, Wang Q, Chen Y, Ni Y, Zhu Y, Joyce BT, Baccarelli A, Deng F, Zhang W, Hou L. Characterization of genome-wide H3K27ac profiles reveals a distinct PM2.5-associated histone modification signature. Environ Health. 2015; 14: 65. doi: 10.1186/S12940-015-0052-5.

33. Liu Q, Ma J, Deng H, Huang SJ, Rao J, Xu WB, Huang JS, Sun SQ, Zhang L. Cardiac-specific methylation patterns of circulating DNA for identification of cardiomyocyte death. BMC Cardiovasc Disord. 2020; 20: 310. doi: 10.1186/s12872-020-01587-x.

34. Lyu Y, Su S, Wang B, Zhu X, Wang X, Zeng EY, Xing B, Tao S. Seasonal and spatial variations in the chemical components and the cellular effects of particulate matter collected in Northern China. Sci Total Environ. 2018; 627: 1627–1637. doi: 10.1016/j.scitotenv.2018.01.224.

35. Mancini FR, Laine JE, Tarallo S, Vlaanderen J, Vermeulen R, van Nunen E, Hoek G, Probst-Hensch N, Imboden M, Jeong A, Gulliver J, Chadeau-Hyam M, Nieuwenhuijsen M, de Kok TM, Piepers J, Krauskopf J, Kleinjans JCS, Vineis P, Naccarati A. microRNA expression profiles and personal monitoring of exposure to particulate matter. Environ Pollut. 2020; 263(Pt B): 114392. doi: 10.1016/j.envpol.2020.114392.

36. Mishra PK, Gorantla VR, Bhargava A, Varshney S, Vashistha P, Maudar KK Molecular detection of Mycobacterium tuberculosis in formalin-fixed, paraffin-embedded tissues and biopsies of gastrointestinal specimens using real-time polymerase chain reaction system. Turk J Gastroenterol. 2010; 21: 129–34.

37. Mishra PK, Raghuram GV, Jain D, Jain SK, Khare NK, Pathak N. Mitochondrial oxidative stress-induced epigenetic modifications in pancreatic epithelial cells. Int J Toxicol. 2014; 33: 116–29. doi: 10.1177/1091581814524064.

38. Morales S, Monzo M, Navarro A. Epigenetic regulation mechanisms of microRNA expression. Biomol Concepts. 2017; 8: 203–212. doi: 10.1515/bmc-2017-0024.

39. Shandilya R, Sobolev AM, Bunkar N, Bhargava A, Goryacheva IY, Mishra PK. Quantum dot nanoconjugates for immuno-detection of circulating cell-free miRNAs. Talanta. 2020; 208:120486. doi: 10.1016/j.talanta.2019.120486.

40. Shen M, Xing J, Ji Q, Li Z, Wang Y, Zhao H, Wang Q, Wang T, Yu L, Zhang X, Sun Y, Zhang Z, Niu Y, Wang H, Chen W, Dai Y, Su W, Duan H. Declining Pulmonary Function in Populations with Long-term Exposure to Polycyclic Aromatic Hydrocarbons-Enriched PM2.5. Environ Sci Technol. 2018; 52: 6610–6616. doi: 10.1021/acs.est.8b00686.

41. Shukla A, Bunkar N, Kumar R, Bhargava A, Tiwari R, Chaudhury K, Goryacheva IY, Mishra PK. Air pollution associated epigenetic modifications: Transgenerational inheritance and underlying molecular mechanisms. Sci Total Environ. 2019; 656: 760–777. doi: 10.1016/j.scitotenv.2018.11.381.

42. STATE OF GLOBAL AIR/2019. A special report on global exposure to air pollution and its disease burden, https://www.stateofglobalair.org/sites/default/files/soga_2019_report.pdf

43. Tessema M, Tassew DD, Yingling CM, Do K, Picchi MA, Wu G, Petersen H, Randell S, Lin Y, Belinsky SA, Tesfaigzi Y. Identification of novel epigenetic abnormalities as sputum biomarkers for lung cancer risk among smokers and COPD patients. Lung Cancer 2020; 146: 189–196. doi: 10.1016/j.lungcan.2020.05.017.

44. Venkataraman C, Brauer M, Tibrewal K, Sadavarte P, Ma Q, Cohen A, Chaliyakunnel S, Frostad J, Klimont Z, Martin RV, Millet DB, Philip S, Walker K, Wang S. Source influence on emission pathways and ambient PM2.5 pollution over India (2015-2050). Atmos Chem Phys. 2018; 18: 8017–8039. doi.org/10.5194/acp-18-8017-2018.

45. WORLD AIR QUALITY REPORT Region & City PM2.5 Ranking. 2019. downloaded from https://www.iqair.com/us/world-most-polluted-cities on 23/07/2020.

46. Xiao T, Ling M, Xu H, Luo F, Xue J, Chen C, Bai J, Zhang Q, Wang Y, Bian Q, Liu Q. NF-κB-regulation of miR-155, via S0CS1/STAT3, is involved in the PM(2.5)-accelerated cell cycle and proliferation of human bronchial epithelial cells. Toxicol Appl Pharmacol. 2019; 377: 114616. doi: 10.1016/j.taap.2019.114616.

47. Yan F, Su L, Chen X, Wang X, Gao H, Zeng Y. Molecular regulation and clinical significance of caveolin-1 methylation in chronic lung diseases. Clin Transl Med. 2020; 10: 151–160. doi: 10.1002/ctm2.2.

48. Yang M, Chu C, Bloom MS, Li S, Chen G, Heinrich J, Markevych I, Knibbs LD, Bowatte G, Dharmage SC, Komppula M, Leskinen A, Hirvonen MR, Roponen M, Jalava P, Wang SQ, Lin S, Zeng XW, Hu LW, Liu KK, Yang BY, Chen W, Guo Y, Dong GH. Is smaller worse? New insights about associations of PM(1) and respiratory health in children and adolescents. Environ Int. 2018; 120: 516–524. doi: 10.1016/j.envint.2018.08.027.

49. Zheng Y, Sanchez-Guerra M, Zhang Z, Joyce BT, Zhong J, Kresovich JK, Liu L, Zhang W, Gao T, Chang D, Osorio-Yanez C, Carmona JJ, Wang S, McCracken JP, Zhang X, Chervona Y, Díaz A, Bertazzi PA, Koutrakis P, Kang CM, Schwartz J, Baccarelli AA, Hou L. Traffic-derived particulate matter exposure and histone H3 modification: A repeated measures study. Environ Res. 2017; 153: 112–119.

